# Genomic inference of sites of transmission during regional spread of *bla_NDM_* ST219 *Klebsiella pneumoniae* in Michigan

**DOI:** 10.1101/2025.10.17.25338245

**Authors:** Tiffany Wan, Sara McNamara, Brenda Brennan, Ali Pirani, Arianna Miles-Jay, Heather Blankenship, Evan S. Snitkin

## Abstract

Carbapenem-resistant *Klebsiella pneumoniae* (CRKP) is recognized as an urgent public health threat due to its multidrug resistance and ability to spread rapidly in healthcare facilities. The frequent movement of colonized and infected patients between different facilities makes it challenging to discern where individual patients acquire CRKP, and in turn, identify facilities making the greatest contributions to regional spread. While high-intensity active surveillance combined with whole-genome sequencing has previously shown success in identifying sites of CRKP transmission, high costs and logistical barriers to regional coordination make this level of surveillance infeasible in most settings. Here, we developed an approach using genomic and healthcare exposure histories from passively collected regional isolates to identify the putative facility source for each new isolate by analyzing shared healthcare exposures with earlier case patients whose isolates shared a most recent common ancestor. As a proof of principle, we applied this approach to data collected by a state health department during a suspected regional CRKP outbreak involving 72 patients exposed to 47 healthcare facilities in Michigan from October 2019 to May 2022. Integration of genomic and healthcare exposure data enabled inference of a single putative source facility for 66/70 of non-index cases, with 35 and 31 cases attributed to a single facility with intra- and inter-facility transmission, respectively. Examination of transmission linkages over time supported a sustained role played by a single focal facility, with several other facilities inferred to be sources of a smaller number of cases. Importantly, in several instances, facilities were implicated as sites of transmission prior to cases being detected there based on overlapping exposures among genomically linked cases, with four facilities being inferred as sites of transmission despite no cases ever being reported. The ability to infer facility sources of transmission using passively collected regional isolates, in some cases ahead of case identification, supports the potential for real-time genome-informed surveillance to enable timely targeting of infection prevention interventions to interrupt regional spread of CRKP and other healthcare-associated pathogens.

## Introduction

Carbapenem-resistant Enterobacterales (CRE) has been recognized as a significant threat to vulnerable patients in healthcare settings, with mortality rates upwards of 50% for patients with CRE infections ^1–6^. Among CREs, New Delhi metallo-β-lactamase (NDM) producing *Klebsiella pneumoniae* outbreaks have been increasingly reported across regional healthcare networks in the U.S.^3,7–9^, with limited treatment options for NDM patients^10^. To prevent NDM-producing strains from reaching levels of endemicity, it is critical to identify and implement more effective strategies to prevent the spread of NDM in healthcare settings.

In addition to many CRE outbreaks that have been investigated at a single facility level ^11–15^, more recent studies have also tracked the spread of CRE across regional healthcare networks ^16–20^. Regional spread presents challenges for individual facilities, as it can be unclear whether a patient newly diagnosed with a CRE infection acquired it during their current hospitalization, or from a facility to which they were previously exposed due to the movement of CRE colonized patients ^21,22^. Knowledge of where CRE transmission is occurring across regional networks is critical to both early intervention for emerging threats like NDM and decreasing rates for endemic organisms ^17,23,24^.

To discern potential facilities of CRE transmission across a region, patients’ histories of healthcare exposure leading up to their clinical CRE culture can be considered ^16–19,25^. In particular, putative sites of acquisition for a given case can be identified by inspecting overlapping exposure with prior cases. However, exposure data alone can be imprecise as circulating cases increase, with frequent overlaps among patients not linked by transmission, and further complicated by unobserved asymptomatic carriers, yielding transmission networks with missing links.

Whole-genome sequencing (WGS) allows for high-resolution delineation of linkage between isolates and has been increasingly applied in regional outbreak investigations ^16–19^. Genomic analysis can mitigate issues with incomplete case capture, as indirect transmission within and between facilities can be inferred using both distance-based or phylogenetic methods. However, identifying transmission from genomic data also has its challenges. Past regional genomic surveillance reports have relied on comprehensive epidemiologic data with high-intensity active surveillance, which is costly and logistically infeasible at regional levels in most settings^26–29^.

Additionally, the current gold standard is to define genetic linkages using genetic distance thresholds that are imprecise, leading to false negative linkages due to intra-patient variation accumulated during prolonged colonization and false positive linkages due to unsampled intermediates ^19,28,30–35^.

In this study, we sought to develop and apply a novel approach integrating WGS and exposure data to identify sites of transmission during a regional outbreak. By leveraging healthcare exposure data and a genetic distance threshold-free approach^12^, within the constraints of a largely passive sample collection with *ad hoc* surveillance conducted at only a few facilities, we sought to identify putative sources of transmission during regional outbreak of *bla*_NDM-1_-carrying *K. pneumoniae* ST219 in Michigan from 2019 to 2022. Taking a facility-centric approach and attempting to discern the origin of each case, we found support for the ability to use sample and data collections commonly available to regional public health labs to identify sites of transmission throughout the outbreak.

## Methods

### Sample collection

Our study was a retrospective analysis of cases of carbapenemase-producing NDM-carrying *K. pneumoniae* in Michigan among hospitalized patients. Cases were reported to the Michigan state public health laboratory and characterized. Specimens were confirmed using Bruker rapifleX® MALDI-TOF, carbapenemase was tested using modified carbapenem inactivation method (mCIM) and carbapenemase genes were identified using the Cepheid Xpert® Carba-R test.

Antimicrobial susceptibility was tested by Sensititre™ GNX2F or GN7F panels. Isolates were subjected to short-read whole genome sequencing using the Illumina MiSeq (See **Supplementary Table 3** for BioSample identifiers and meta-data). Healthcare exposures in the 90 days preceding first case detection of a given patient was also collected using the state electronic surveillance system.

### Genomic data processing

To identify previously sequenced strains that were closely related to outbreak isolates, annotated public genome assemblies listed as “*Klebsiella pneumoniae”* were downloaded from the PATRIC database ^36^, samples were annotated by RASTtk ^37^, and a core genome distance matrix was generated using the R package cognac ^38^. We then selected public genomes with core genome distances within 200 single nucleotide variants (SNVs) of at least one of the outbreak genomes. Antibiotic resistant genes in outbreak and public genomes were identified using AMRFinderPlus v3.11 ^39^.

For tracking the spread of outbreak strains, we performed variant calling using a custom reference-based pipeline (https://github.com/Snitkin-Lab-Umich/snpkit). The quality of sequencing reads was assessed using FastQC v0.11.0, and adapter sequences and low-quality bases removed using Trimmomatic v0.39. Single nucleotide variants (SNVs) were identified by first using Burrows-Wheeler short-read aligner (bwa v0.7.17) to map trimmed reads to the ST219 reference genome (SAMN26729713), then discarding polymerase chain reaction (PCR) duplicates with Picard v3.0.0, and calling variants with SAMtools and bcftools v1.9. Variants were filtered from using VariantFiltration from GATK v4.5.0.0 (QUAL > 100; MQ>50;>=10 reads supporting variant; and FQ< 0.025). We performed GATK HaplotypeCaller for indel calling only including those with root mean square quality (MQ) > 50.0, GATK QualbyDepth (QD) > 2.0, read depth (DP) > 9.0, and allele frequency (AF) > 0.9. We also excluded variants that were less than 5 base pairs in the proximity to indels, in recombinant regions identified by Gubbins v3.0.0, in a phage region identified by Phaster web tool, or resided in tandem repeats of length greater than 20 bp as determined using the exact-tandem program in MuMmer v3.23 using a custom Python script. The whole-genome masked variant alignment generated by the variant calling pipeline was then used to reconstruct a maximum likelihood phylogeny with IQ-TREE v2.0.3 using the general time reversible model GTR+G and ultrafasta bootstrap with 1000 replicates (-bb 1000). To estimate common ancestors of collected isolates, a dated phylogeny was generated using Bactdating “arc” model was chosen due to the lowest DIC and smallest root date confidence interval (**Table S1**) ^40^

### Identifying epidemiologic and genomic links between patients

All facilities a patient was exposed to prior to isolation of NDM *K. pneumoniae* were considered as potential sources of acquisition. For the facility where the isolate was collected, it was required that the collection occurred more than 2 days after admission for that facility to be considered a potential source. The exception to this was for cases where there were no documented healthcare exposures prior to the facility of isolation, in which case it was considered as a potential source. In identifying genetic and epidemiologic support for a case’s facility source of acquisition, all prior cases were considered putative sources, with the patient’s shared history of exposure to healthcare facilities used to provide epidemiologic support. We did not require temporal overlap between cases for establishing linkages, to enable consideration of indirect transmission via environmental reservoirs or undetected patient intermediates.

To simulate tracking of transmission in real-time we identified the putative healthcare facility source of each new case patient’s strain by considering isolate genomes and exposure data from patients with earlier isolate collection dates, in other words, we considered exposure data that would be available at the time patients tested positive. Our approach consisted of first identifying prior isolates with whom a case isolate shares a most recent common ancestor as inferred by maximum shared variants (MSVs) and then examining patterns of shared healthcare exposure for MSV pairs. If a case isolate had MSVs with multiple isolates, then all potential pairs were considered. To identify putative source facilities, five different shared healthcare exposure types were extracted (**Figure S1a**): (i) if both the case patient and the MSV linked patient had their isolate collected at the same facility, then this was classified as putative intra-facility transmission at the site of collection, (ii) if the MSV linked patient had prior exposure to the facility where the case patient’s isolate was collected, then this was classified as a putative intra-facility transmission at the case patient collection facility, (iii) if the case patient had prior exposure to the facility where the MSV-linked patient had their isolate collected, then this facility was inferred as the source, (iv) if the case patient and the MSV-linked patient shared exposure to a common third facility, then this facility was inferred as the source, and (v) if there were no shared healthcare exposures between a case patient and their MSV-linked patient, then the source facility was inferred as the facility where the MSV-linked patient had their isolate collected.

For instances where a case patient was linked to more than one patient based on MSVs, all linked patients were considered. However, to focus on stronger epidemiologic hypotheses, we added an optional filter to prioritize facilities implicated by the majority of MSV-linked patients. Using the filter, for a given case patient that shared healthcare exposures with multiple MSV-linked patients and greater than 50% of putative source facility attributions based on criteria (i)-(iv) above were to the same facility, then the single facility was deemed the putative source with both genomic and epidemiologic support. Similarly, if a given case patient had no shared healthcare exposures with MSV-linked patients (criteria (v) above), but more than 50% of isolates from MSV-linked patients were collected at the same facility, then the facility was deemed the single putative source facility with genomic support.

### Evaluation of putative transmission linkages

Several strategies were used to evaluate the veracity of putative transmission events. First, we evaluated whether using MSVs was linking patients with shared healthcare exposure history more than expected by random. To this end we performed a permutation analysis wherein each patient’s association with their isolate was maintained, but their history of healthcare exposures was randomly swapped with another patient. We compared the proportion of MSV linked pairs that had no shared healthcare exposure of our data with the 1000 permutated events using Z-test. Second, we evaluated the patient-sharing of facilities with predicted inter-facility transmissions between them. To accomplish this, we used inpatient billings from Center of Medicare and Medicaid services (CMS) and inferred patient transfer frequencies between facilities based on sequential billings. Using the raw patient transfer matrix for CMS patients, we calculated the patient flow between pairs of facilities using regentrans ^41^. For each case patient, we compared the patient flow between the case facility and predicted source facilities, to (i) patient flow between case facility and a facility where case was previously exposed to but not predicted as a source facility, and (ii) the flow between the case facility and other facilities that were shared patients in the CMS data, but were not predicted as linked to the case facility. We used Wilcoxin-signed rank test to compare the patient flows. Lastly, we evaluated the importance of each facility at time points when cases were collected using eigencentrality and pagerank algorithms **(Figure S4)**.

### Statistical analysis

All statistical analysis were done in R. Network algorithms were implemented using igraph^42^ in R. Figures were visualized in R using ggplot2 ^43^, ggtree ^44^ and ggraph (https://github.com/thomasp85/ggraph).

## Results

### Regional spread of NDM-harboring *K. pneumoniae* ST219 was attributed to clonal expansion

From October 2019 to April 2022, 104 *Klebsiella pneumoniae* ST219 isolates harboring the *bla_NDM-1_* gene were collected from 72 individuals across regional healthcare facilities in Michigan. For the first six months, cases were only recovered at a single acute care hospital (ACH10). During the next phase of the outbreak, cases were reported by 21 additional healthcare facilities, with the peak occurring from June 2020 to June 2021 **(Figure 1a)**. Isolates were compared using whole-genome sequencing to understand whether the shift from a putative single facility outbreak to a broader regional spread was due to a single regional introduction or multiple introductions of NDM-carrying *K. pneumoniae*. Phylogenetic reconstruction of Michigan isolates, along with publicly available *K. pneumoniae* ST219 genomes showed Michigan isolates separate from public isolates in a distinct monophyletic cluster, consistent with a single introduction into the region **(Figure S2)**. Further support for all cases deriving from a single recent introduction came from the small genetic distances among Michigan isolates, with a median of 8 pairwise SNVs (36 maximum, 0 minimum) (**Figure S3**). Of note, public *K. pneumoniae* ST219 genomes from diverse geographical locations showed variable association with *bla_NDM-1_*, indicating that *bla*_NDM-1_ gene had been acquired multiple times in this genetic background (**Figure S2**).

**Figure 1.**
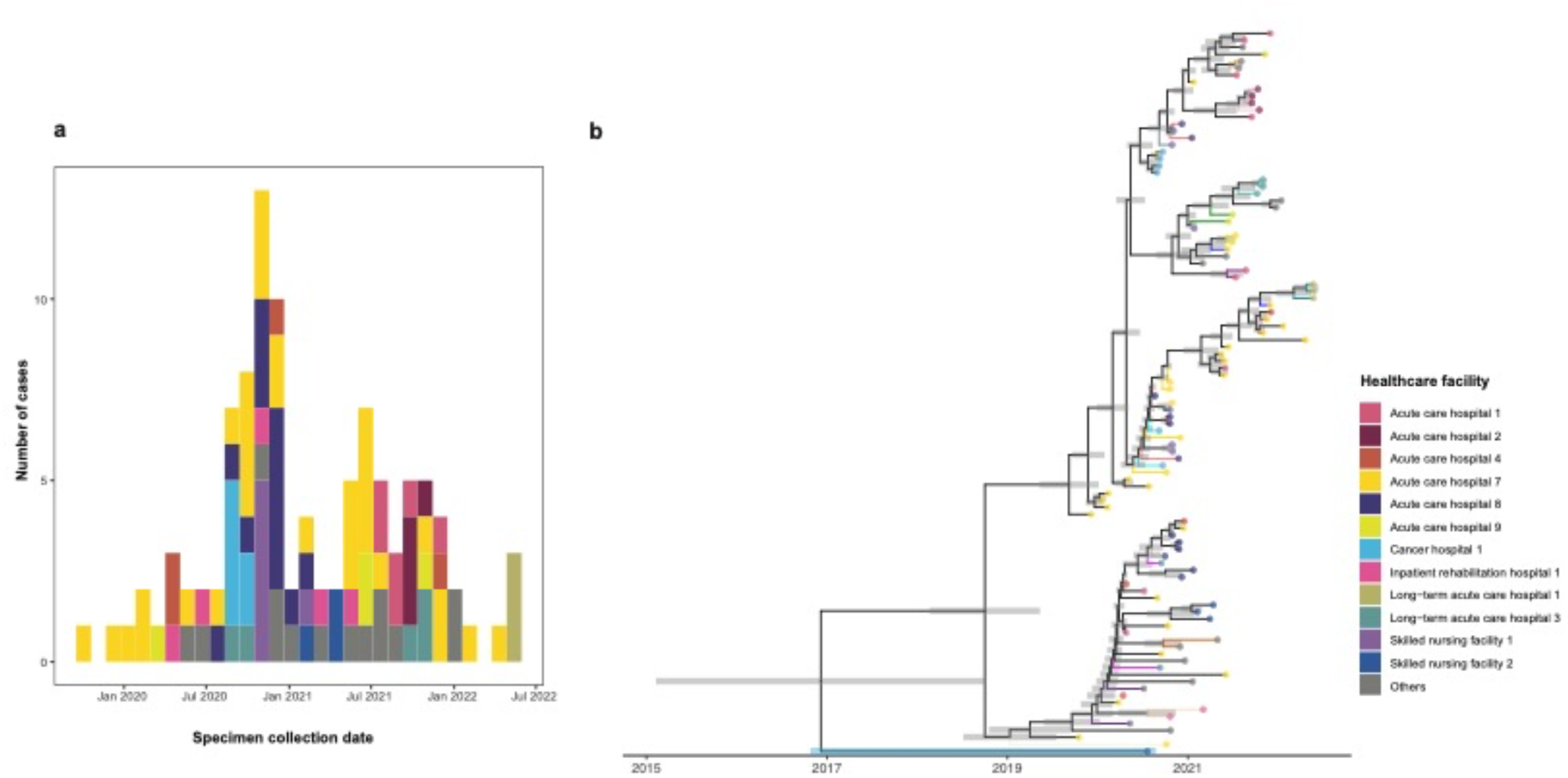
Epidemiologic and genetic description of *bla_NDM-1_*-carrying ST219 *K. pneumoniae* outbreak strains from Michigan. **(a)** Epidemic curve showing number of *bla_NDM-1_* ST219 cases collected from different Michigan healthcare facilities from 2019 to 2022. **(b)** Time-calibrated phylogeny of outbreak genomes with gray bars indicating the 95% confidence interval of ancestral date. Edges are the same color if from the same patient and black otherwise. Tips are colored by healthcare facility of collection, with gray representing all facilities with less than 3 cases identified. The outlier isolate with a common-ancestor predating the outbreak is highlighted in light blue (See main text).

To further understand the temporal dynamics of clonal spread, we created a time-scaled phylogeny of Michigan isolates (**Figure 1b**). The time-scaled phylogenetic tree of Michigan isolates was also consistent with clonal spread, with the common ancestor of all but one isolate estimated to be in early 2019 (Confidence interval (CI): 2018.271, 2019.495). Although detected in 2020, the one outlier isolate had a predicted common ancestor with the other outbreak isolates that dated back to 2017 (CI: 2015.15, 2018.81). We noted that this outlier isolate was collected at a facility where cases from the dominant outbreak lineage were detected contemporaneously, consistent with independent acquisition of the *bla_NDM-1_*-encoding plasmid by a closely related *K. pneumoniae* ST219 strain. Combined with the observation that the most closely related public isolate to the Michigan outbreak lacked *bla_NDM-1_*, this supports the outbreak clone stemming from the acquisition of *bla_NDM-1_* in a locally circulating strain of ST219.

### Patients with genetically related isolates showed extensive overlap at regional healthcare facilities

To understand the pathways by which the major outbreak lineage spread within and between healthcare facilities, we overlaid a whole-genome maximum-likelihood phylogeny with patients’ healthcare transfer histories **(Figures 2a and 2b)**. Visual inspection of **Figure 2** showed numerous instances of temporal overlap in healthcare exposures among patients whose isolates clustered on the whole-genome phylogeny. These overlaps supported cases of intra-facility transmission (i.e. phylogenetically clustered cases detected at the same facility), direct inter-facility transmission (i.e. phylogenetically clustered cases with common exposure to one of the patient’s collection facilities), and indirect inter-facility transmission (i.e. phylogenetically clustered cases with common exposure to a third facility, distinct from the facilities where patient’s isolates were collected). Moreover, overlaying the phylogeny with healthcare exposures also revealed instances where cases detected contemporaneously at the same facility were clearly due to independent importation events. For example, in late 2020, strains collected concurrently through active surveillance at ACH10, were from two distinct genomic clusters sharing a more distant common ancestor.

**Figure 2.**
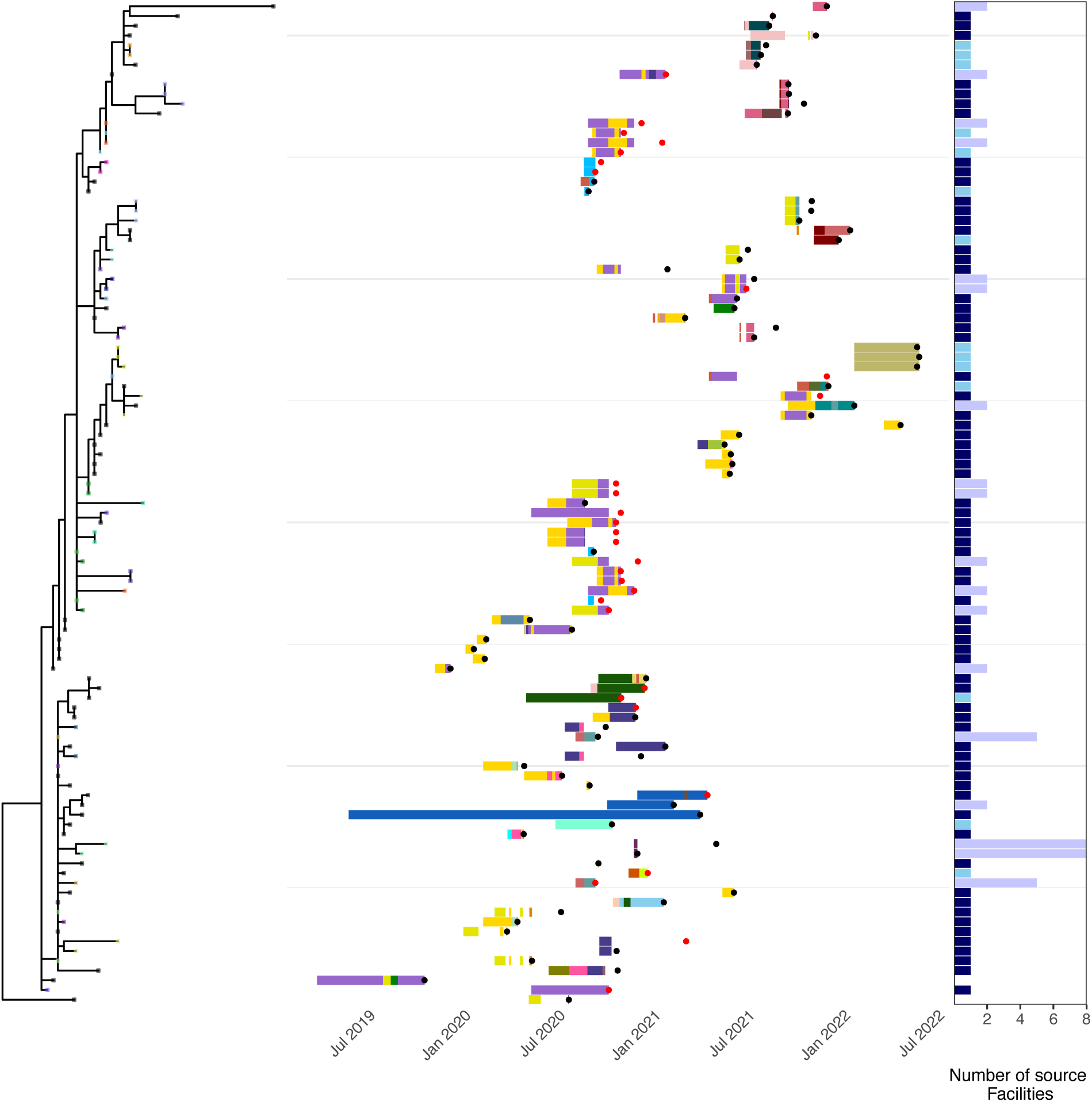
Whole-genome phylogeny of outbreak isolates overlaid with associated patient’s history of healthcare exposures. **(a)** A whole-genome maximum-likelihood phylogeny is shown for outbreak ST219 strains. Tips of the phylogeny are colored for patients with multiple isolates, with each color indicating a different patient or black if patient only had one isolate. **(b)** The history of patient’s healthcare facility exposures leading up to their isolate collection are shown for patients with isolates represented on the phylogeny in panel **a**. The x-axis indicates dates, different color bars indicate stays in different healthcare facilities, black circles indicate a collected clinical isolate and red circles represent isolates collected via active surveillance. **(c)** The source facility(ies) for each isolate was predicted based on shared healthcare exposures with other patient(s) with whom their isolates shared MSVs. Bars with a height of one indicate >50% of links support that facility, with higher values indicating multiple linked source facilities. Colors indicate whether links were supported by genomic and epidemiologic data (dark blue), genomic data only (light blue), or a single facility not supported (purple).

### Integrating genomic and healthcare exposure data revealed regional transmission pathways

Motivated by the overlapping healthcare exposures among phylogenetic neighbors, we sought to systematically extract intra-facility and inter-facility transmission events by considering both genomic and healthcare exposure data. To this end, we simulated the tracking of the NDM-carrying *K. pneumoniae* ST219 regional spread as a real-time event, by inferring transmission events based on linkages between “new cases” and cases identified prior. The putative source(s) for each case were inferred to be previously collected isolates with which their isolate had maximum shared variants (MSV) (**Figure S1a**). In contrast to the common approach of employing an SNV threshold to detect transmission linkages, MSV pair detection did not rely on a single threshold, rather identifying circulating strains with which they share a most recent common ancestor. Among pairs with maximum shared variants there was a median of 3 cgSNVs (IQR:1,5), while also permissive to larger variance (maximum: 22 cgSNVs, **Figure 3a**).

**Figure 3.**
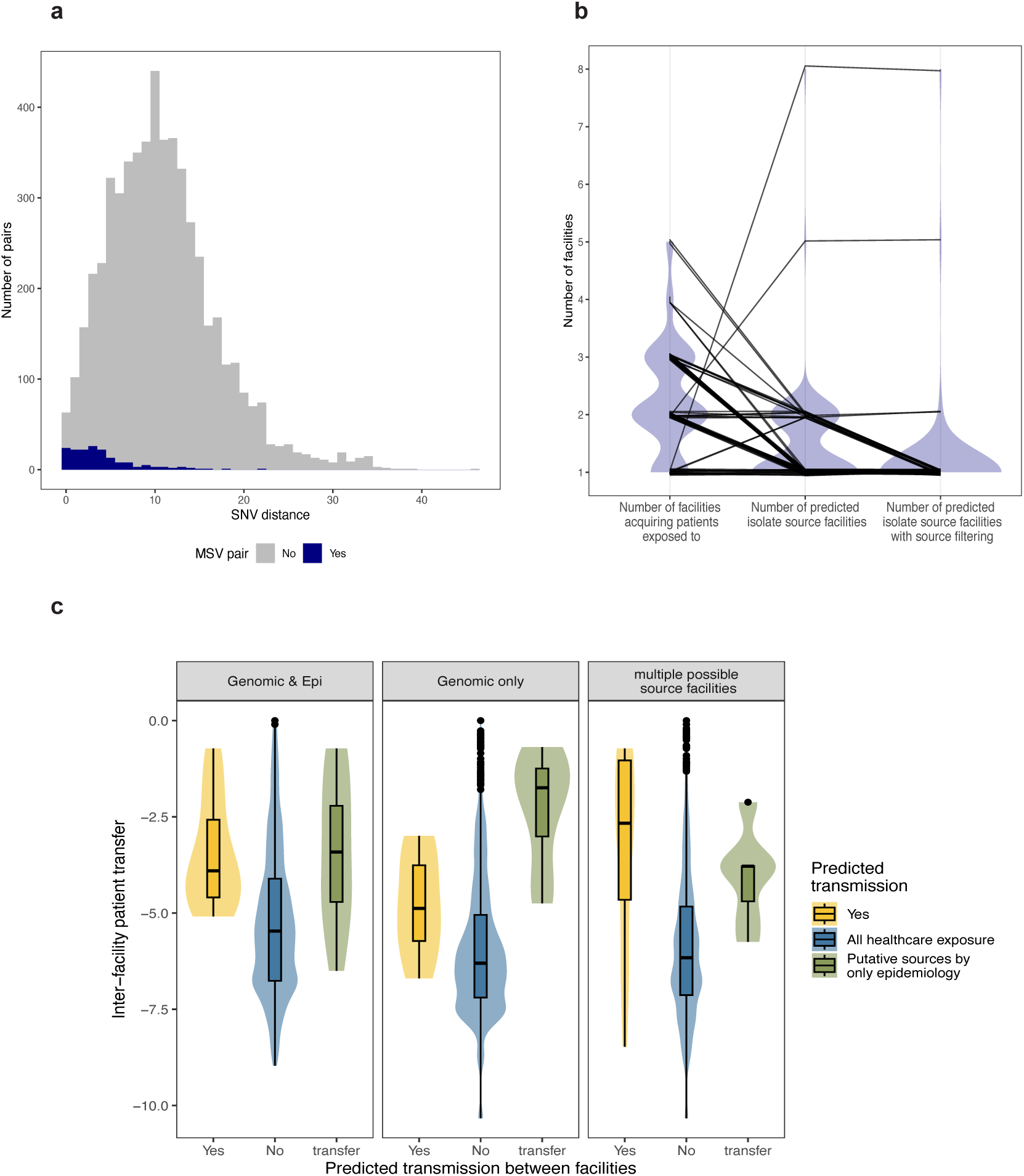
Evaluation of transmission linkages inferred using maximum shared variants (MSVs). **(a)** Histogram of pairwise distances for all pairs of isolates collected during outbreak (gray), and MSV pairs (blue). **(b)** Spaghetti plot comparing the number of healthcare exposures for each patient (left), the number of putative source facilities based on genomic and/or epidemiologic links to MSV pairs (middle) and the number of putative source facilities when selecting a single facility if it is represented by >50% of exposures with MSV pairs (right). **(c)** Statewide patient transfer networks derived from CMS billing data were used to calculate the magnitude of the patient flow between all pairs of facilities from which case patients were identified. Patient flow in log scale between source and case facilities with predicted transmissions (yellow), from all facilities connected to the case facility in the CMS network(blue) and from all facilities the case patient was exposed to the case facility (green) are shown for cases where the putative source facility had genomic and epidemiologic support (left), genomic support only (middle) and cases where no single source facility was supported by >50% of links (right) .

Using this simulated real-time approach, we identified 206 candidate source-to-case links among the 70 isolates included in the analysis, where cases could be linked to more than one source if they had MSV with more than one previously detected isolate. To determine pathways of intra- and inter-facility transmission, patterns of healthcare overlap were analyzed among MSV pairs. Our objective was to identify the most plausible facility where each case acquired their isolate, therefore, we assigned a single high-confidence source facility to each case when more than 50% of links to a case supported the same source facility (**Figure S1b**). Using this approach, 50% (35/70) of patients were predicted to acquire their isolate via intra-facility transmission events at 9 different facilities. Among the 35 putative intra-facility transmissions, 6 patients also had potential inter-facility transmission with shared exposure with another healthcare facility. The remaining 35 cases were attributed to inter-facility transmissions, with 60.0% (21/35) supported by both genomic and shared healthcare exposure, 28.6% (10/35) supported only by genomic linkages and 11.4% (4/35) of inter-facility cases assigned to more than one facility meeting the high-confidence source facility criteria (**Figure 2c**). These results show that by combining MSVs with healthcare exposure data we were able to effectively narrow down the putative source facilities, with 94.3% (66/70) of patients having a single predicted source facility for their isolate (**Figure 3b**).

### Predicted transmissions are enriched in shared healthcare exposures and link facilities that share more patients than unlinked facilities

To evaluate the transmissions predicted using our approach, we conducted two additional analyses. First, we performed permutation analysis to evaluate whether healthcare-sharing among genomically linked pairs occurred more than expected. We created 1000 randomized data sets, where patients were assigned another patient’s history of healthcare exposures. In none of these permuted data sets were there more instances of shared healthcare exposure among MSV pairs than in the actual data (p < 0.001). This result supports the MSV approach identifying patients with shared exposures far more than expected by chance (**Figure S6)**.

Next, we assessed support specifically for predicted inter-facility transmissions. To provide context to our predictions, we leveraged statewide patient transfer networks inferred by sequential billing events extracted from Centers for Medicare and Medicaid databases (CMS), which provides a quantitative summary of how patients move between regional healthcare facilities. We first focused on predicted transmissions with both genomic and epidemiologic support. For this set of predictions, we observed that source and case facilities linked by transmission had significantly higher patient flow with each other than did other facilities connected to the case facility in the CMS network (**Figure 3c**, p = 2.54 × 10^−9^), indicating that transmissions are predicted to preferentially occur between facilities that share the most patients. However, putative source facilities in linkages supported by genomic and epidemiologic data were did not have significantly higher patient flow to the case facility than the other facilities that the case patients were exposed to (**Figure 3c, p = 0.89**). This observation indicates that the genomic data can help differentiate among alternative epidemiologically plausible scenarios by helping identify the most likely source facility among multiple past exposures for a given case patient (**Figure 3c, Supplemental Table 2**).

Next, we examined predicted inter-facility transmissions supported only by genomic data. As with predictions with epidemiologic support, facilities with genomic-only transmissions had significantly higher patient flow than other facilities linked to the case facility in the CMS network (**Figure 3c**, p = 0.02). However, source and case facilities linked by transmission with only genomic support had significantly lower patient flow to the case facility than the other facilities case patients in these linkages were exposed to (**Figure 3c**, p =0.0001). Thus, genomic only prediction are linking facilities with an intermediate level of patient sharing. To further assess the plausibility of these linkages, we examined the genomic support more closely. Overall, the putative transmission pairs with only genomic support were highly clonal (IQR [1.5,5], min = 0, max = 12). Moreover, for four cases with healthcare overlap with other putative source patients without MSVs that were collected prior to the cases, the pairwise SNVs were significantly larger (IQR = (5.75,10.25), min =4, max = 18) compared to the genomically linked putative source (IQR = (0.75,3.25), min = 0, max =4). In addition, two of the ten patients with only genomic evidence were linked to a single source facility by more than one source patient, further supporting connectivity between the facilities despite the lack of direct healthcare exposure.

### Regional genomic analysis enables identification of critical facilities over time

Having developed and validated an approach to track intra- and inter-facility transmission, we next examined patterns of transmission over the course of the outbreak at both patient- (**Figure 4a**) and healthcare facility-level (**Figure 4b**). Overall, we identified 64 independent inter-facility transmissions. ACH10, where the outbreak was first detected, was predicted to be a persistent site of both intra- and inter-facility transmission throughout the study period. With respect to inter-facility transmission, ACH10 was predicted to be a source for 26.6% (17/64) of inter-facility transmission events and the destination for 15.6% (10/64) (**Figure 4a**). Aside from links involving ACH10, most other inter-facility links were only observed once, with at most a small number of subsequent intra-facility transmissions detected after the introduction. Moreover, when considering exportations from facilities in aggregate and using the eigencentrality and pagerank algorithms ^42^ to calculate facility importance, ACH10 was consistently the most important facility throughout the outbreak (**Figure 5** and **Figure S5**).

**Figure 4.**
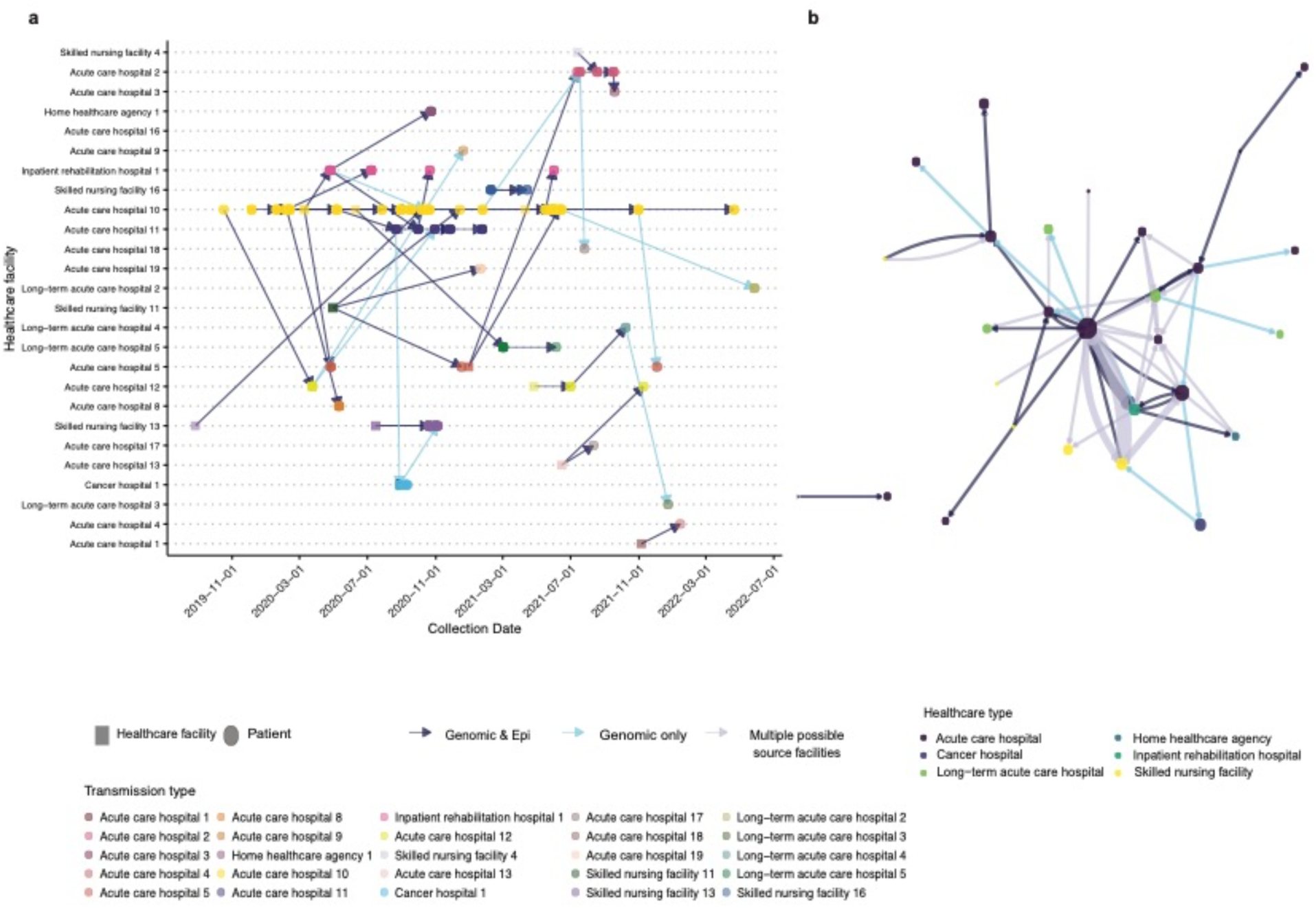
Inferred patient- and facility-level transmission networks. **(a)** For the patient-level network, each node represents a different patient (circles) or a facility predicted to mediate indirect transmission (triangles). The x-axis is time and the y-axis is the healthcare facility where an isolate was collected. Healthcare facilities are also represented with color to enable easier visual tracking horizontally. Arrows are drawn between patients with genomic only (black), or genomic plus epidemiologic linkage (blue). The direction of the arrows between two patients indicates the direction of intra- or inter-facility transmission supported by epidemiologic data. **(b)** For the facility-level network, each node represents a different healthcare facility colored by facility type. The size of the node corresponds to the number of cases isolated from the facility and nodes. The arrows connecting two nodes are the predicted inter-facility transmissions and the width of the edges are the number of predicted transmissions between a pair of facilities. The colors of the arrows indicate whether the predicted linkage was genomic and epidemiologically supported (navy blue), only genomically supported (sky blue), or having multiple possible sources (gray blue).

**Figure 5.**
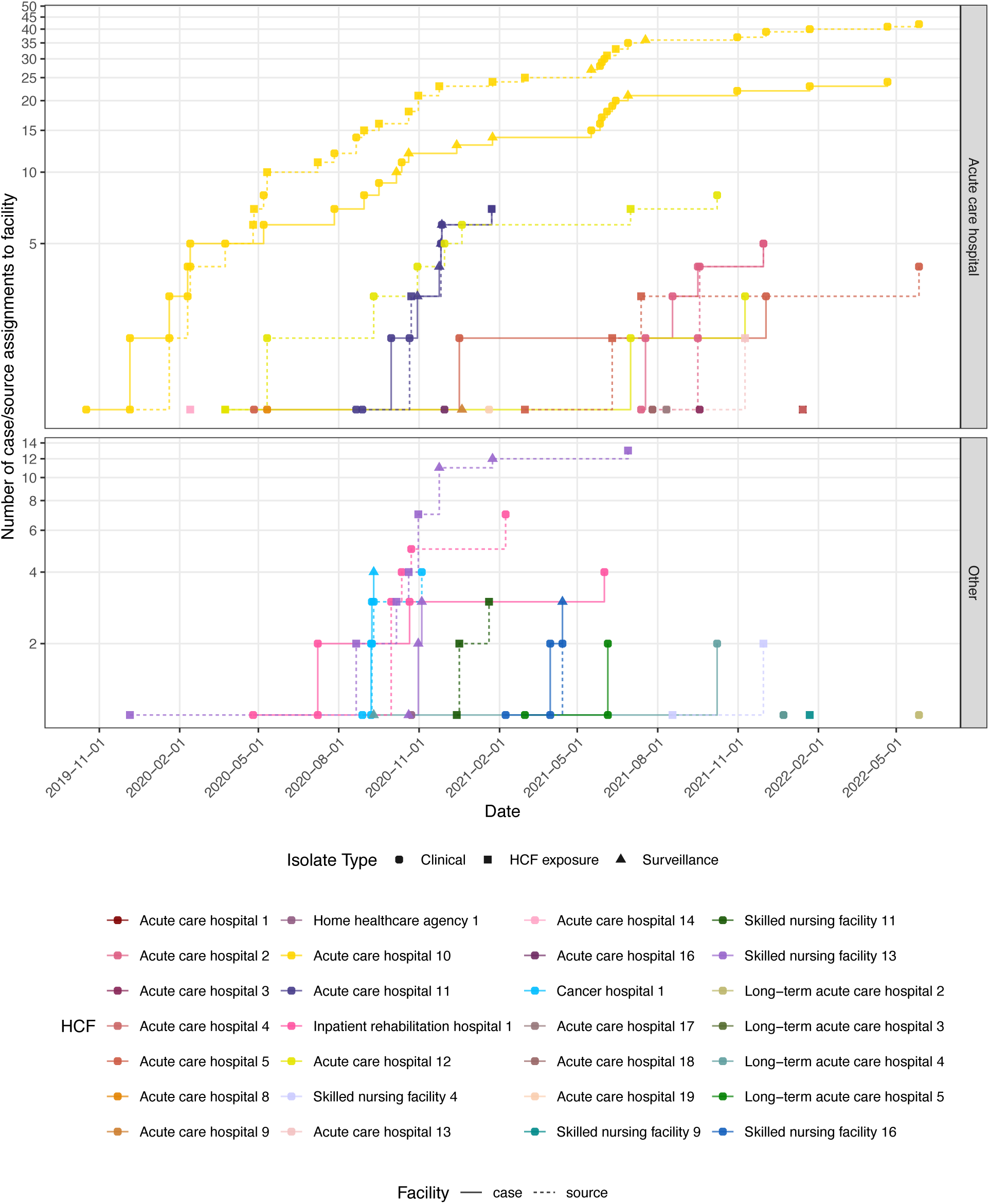
Healthcare facility’s predicted roles in outbreak over time. The cumulative case and source curves for each healthcare facility are shown over time upon new case identification. Here, acute care hospitals (top) are shown separately from the other healthcare types (bottom). Each new case increases the case curve by one in the facility of isolation, and in turn increases the putative source curve for either the same healthcare facility (predicted intra-facility transmission) or another healthcare facility (predicted inter-facility transmission). Circles indicate clinical isolates, squares indicate surveillance isolates, and triangles indicate sources inferred through indirect inter-facility transmission.

While analyses of the transmission network indicated that ACH10 was an important player throughout the outbreak, we were interested to see if there were more contributions from other facilities over the course of the outbreak. To this end, we examined the number of cases sourced to each facility over time. By comparing facility-level source curves to facility-level case curves, we were able to discern predicted intra-facility transmissions, importations and exportations from each facility over time (**Figure 5**). For example, by examining ACH10’s source and case curve, we were able to see the transition from initial predominance of intra-facility transmissions (source curve and case curve rising in sync) to exportation (source curve increases, while case curve remains flat). For most other facilities we see the case curve initially rises, indicating importation, followed by increases in the source and case curve during subsequent periods of intra-facility transmission and/or exportation (e.g. facilities ACH2, ACH5 and ACH11).

However, we noticed some facilities with unexpected patterns, whereby they were inferred as sources of transmission prior to any cases being detected. For SNF13, we observed the source curve rising before any cases were detected, with its role as a source being inferred based upon shared exposure of MSV pairs from other facilities. In total, there were three cases with genomic and epidemiologic support for SNF13 being a source, with two of these cases occurring before the first case was identified in SNF13. It is worth noting that while no cases were initially identified at SNF13, patients from ACH10 were often transferred from SNF13, supporting its potential role as an undetected reservoir early in the outbreak. In addition to SNF13, there were four other facilities with no cases ever reported, but for which shared exposures implicated them as a potential source of regional transmission. Support for cases being sourced to these facilities which never reported cases comes from many of these putative transmission pairs having extremely small genetic distances, consistent with recent shared exposures mediating transmission. Case NDM165 attributed to source facility ACH1 had 0 SNVs with its predicted source isolate. Case NDM130 and case NDM150 both had a putative source with 3 SNVs sharing healthcare exposure at ACH13. Case NDM89, NDM90, and NDM94 had 0 SNVs, 3 SNVs and 0 SNVs with shared health exposure at SNF11 respectively. Lastly, case NDM132 and case NDM156 were sourced at SNF4, with 6 SNVs and 20 SNVs with their respective putative source.

## Discussion

Healthcare-associated infections caused by antibiotic resistant organisms are a major public health threat, associated with significant morbidity, mortality and economic impacts^45,46^. Most efforts to date to track and prevent the spread of infections have focused on individual healthcare settings. However, it is increasingly appreciated that a regional perspective is critical to identify the sites of transmission and pathways of inter-facility spread that initiate and sustain regional endemicity ^26–29^. While regional active surveillance combined with genomic analysis has been shown to enable regional transmission tracking in low prevalence regions, this strategy is costly and typically infeasible to implement at the state or province level. Here, we implemented and evaluated a regional approach using genomic and healthcare exposure data to identify putative source facilities where patients may have acquired their infection, without relying on an established active surveillance program. Applying this approach to a *bla_NDM-1_* carrying *K. pneumoniae* ST219 regional outbreak in Michigan, we identified putative facility sources of patient’s isolates with high specificity, identified key facilities seeding cases in other facilities and showed the potential to identify facilities as sources of infection prior to their detection of cases. These findings support the potential for regional genomic surveillance programs using passive isolate collections and ad hoc surveillance to guide regional infection prevention action to target facilities driving regional spread.

Mapping patient transfer history on the whole-genome phylogeny of patients’ acquired strains showed that phylogenetically clustered patients tend to share healthcare exposure history. This observation, along with the success of prior studies using phylogeographic methods to track regional healthcare transmission ^18,28,47–50^, prompted us to use a maximal shared variant approach to infer the facility source of each patient’s isolate. The MSV approach leverages common ancestry but does not require phylogenetic reconstruction when a new isolate is sequenced, will scale to large numbers of strains, and is easily implementable and interpretable. Compared to the standard approach of imposing a genetic distance cutoff, MSV genomic links yielded isolate pairs with SNV distances comparable to those reported in prior *K. pneumoniae* transmission studies ^19,28,30–35,51^, while also allowing for larger genetic distances that may arise due to prolonged patient colonization and hypermutators^12^. Here, we showed that the MSV approach yielded links among patients that were significantly enriched in shared healthcare exposures, identified connections among facilities that are highly connected by patient transfer, and provided sufficient resolution to track the spread of a regional CRE outbreak.

Upon identification of MSV pairs, we implemented an approach to incorporate facility exposure data with an eye towards real-time surveillance, that focused on elucidating the contribution of individual facilities to regional spread. To simulate a real-time response, for each new case we only considered earlier cases when identifying putative source facilities. While imperfect, as a true source may only become apparent later due to the lack of active surveillance, only retrospective case data would be available in practice. To increase robustness to missing case capture caused by delayed case reporting and limitations in surveillance, we did not require temporal overlap in hospitalizations of genomically linked patients, but rather just that there was shared exposure prior to a case patient’s isolate collection. In this way, we could capture intra- or inter-facility transmission that may be mediated by one or more unsampled intermediate patients at the facility of interest. The viability of this approach was demonstrated by both the non-random concordance between genomic linkages and exposure data, as well as the specificity of predictions, with 94% (66/70) of patients being linked to one putative source facility. We note that to achieve this specificity, when greater than 50% of the shared exposures for a case patient with other patients linked by MSVs were from a single facility, this single facility was inferred to be the source. This specificity filter simplified the network and prioritized scenarios most supported by genomic and exposure data, aligning with our motivation to prioritize facilities for intervention, but in practice all putative source facilities could be retained and investigated. For the four cases with multiple putative source facilities after imposition of the specificity filter, this was often due to the frequent movement of patients back and forth among a common set of facilities. Thus, although a specific source facility was not identified in these cases, a cluster of connected facilities putatively exposed to carriers was identified.

Analyzing the predicted patterns of regional transmission over the course of the outbreak revealed a single facility playing a central role. ACH10 was the first facility with cases identified, had the highest number of predicted intra-facility transmissions, and was predicted as the most common source of transmissions to other facilities. Our findings were concordant with previous studies of regional CRE outbreaks, which have consistently observed a small number of focal facilities either seeding initial regional spread, or sustaining it over time ^17,18,48^. In past reports where focal facilities were long-term care facilities, authors speculated that uncontrolled transmission at these facilities was critical for establishing a reservoir that subsequently spread to connected facilities ^17,18^. However, in cases where the focal facility is an ACH that shares patients with many other facilities, it is possible that one or more of these connected facilities is acting as a potentially unsampled reservoir ^48^ For example, patients from SNF13 were often later transferred to ACH10 in our study. Alternatively, the sustained presence of a strain at a single facility could be due to environmental or plumbing contamination, periodically seeding small clusters ^52^. Regardless, both this and prior studies show the potential of genomic surveillance to hone in on key facilities and help prioritize surveillance and intervention activities. In addition to identifying focal facilities, we also noted that integration of genomic and exposure data enabled early detection of facilities contributing to regional spread. This manifested in both the inference of facilities as sources of infections in other facilities prior to the source facility detecting cases, as well as inferring facilities as sources that never reported cases. This demonstrates the power of this integrated approach to help identify potential key facilities early in an outbreak before high case counts are reported.

Our study has several important limitations to consider. First, most facilities only contributed clinical isolates, with only a few facilities performing ad hoc surveillance to detect NDM carriers. For example, ACH7, had the most cases and performed the most surveillance over the study period. Given the known iceberg effect whereby asymptomatic carriers greatly outnumber patients with overt infection ^4,53–57^, we likely missed many cases due to this passive approach.

However, widespread active surveillance is likely to be infeasible in most settings. Therefore, we implemented our analysis to be focused on tracking routes of transmission within and between facilities, as opposed to constructing patient-level transmission networks. Here we show potential to support use genomic analysis to guide targeted active surveillance efforts, rather than regional active surveillance, to more efficiently utilize limited resources. The robustness of our approach was supported by non-random healthcare exposure overlap among genetically linked cases, as well as facilities with genomic linkages sharing significantly more patients’ according to CMS claims data. A second limitation is that we only had access to healthcare exposure information up until patients became a case (i.e. until *bla_NDM-1_*-carrying case was detected via surveillance or clinical culture). Therefore, we may be missing shared healthcare exposures that took place after case identification. However, despite this missing data, we were able to capture transmission linkages for most cases with high specificity, indicating that sufficient exposure data was available to link cases between facilities. Additionally, we only had healthcare exposure 90 days prior to CRE detection, while previous studies suggested CRE can colonize patients for months and years ^58^. Therefore, a longer period of healthcare exposure data may further inform inter-facility transmission. Third, our analysis focused on a region where NDM is not yet endemic, with cases likely originating from a single introduction and clonal spread, which may limit generalizability. Demonstrating the clonality of the outbreak enabled us to use the MSV approach without additional filtering by genetic distance. However, in a more complex setting including frequent outside importation to the region and/or plasmid transfer, distance thresholds and/or regional context may need to be considered.

Overall, we implemented a threshold-free approach to collate genomic and healthcare exposure data and applied it to elucidate the role of healthcare facilities as sources of transmission during a CRE outbreak in Michigan. Our approach has the potential to shed light on important healthcare facilities mediating transmission and capture missed facilities early on during the outbreak. As shown in previous studies, our results highlighted the value of using patient transfer history rather than solely relying on sites of case identification. By enabling early detection of key facilities of the outbreak, our approach could guide selection of facilities for enhanced genomic surveillance throughout the outbreak to prevent the spread of antibiotic resistance.

## Data Availability

All whole genome sequence data are available online at NCBI
All code used for data analysis are available online at github

https://github.com/tifwan/ST219_NDM_ms

---

**Supplementary Table 1.** Bactdating model statistics.

| Model | $\mu$ ESS | $\sigma$ ESS | $\alpha$ ESS | Clock rate | Root date | Root date lower CI | Root date upper CI | DIC |
| --- | --- | --- | --- | --- | --- | --- | --- | --- |
| poisson | 65.03 | 0 | 501 | 417.55 | 2016.58 | 2016.45 | 2016.69 | 94542.89 |
| negbin | 571.75 | 501 | 393.63 | 855.32 | 2013.03 | 2002.72 | 2017.79 | 1688.33 |
| strictgamma | 1.82 | 0 | 15.06 | 117.27 | 2008.71 | 2007.08 | 2010.29 | 289820.68 |
| relaxedgamma | 501 | 501 | 501 | 860.42 | 2013.5 | 2004.91 | 2017.7 | 1724.12 |
| mixedgamma | 414.48 | 411.74 | 427.9 | 854.09 | 2013.13 | 2004.34 | 2017.98 | 1727.73 |
| arc | 501 | 698.44 | 501 | 774.06 | 2017.37 | 2015.15 | 2018.81 | 1671.2 |
| carc | 532.6 | 469.23 | 426.1 | 747.82 | 2016.98 | 2014.57 | 2018.64 | 1726.6 |
| mixedcarc | 866.45 | 1196.54 | 501 | 748.4 | 2016.93 | 2014.16 | 2018.66 | 1689.79 |

**Supplementary Table 2.** Confidence interval and significance level of CMS evaluation.

| Group 1 | Group 2 | Genomic and Epi |  |  |  | Genomic only |  |  |  | Multiple possible source facility |  |  |  |
| --- | --- | --- | --- | --- | --- | --- | --- | --- | --- | --- | --- | --- | --- |
|  |  | Estimate [CI] | p-value | W | # Group1/<br># Group2 | Estimate [CI] | p-value | W | # Group1/<br># Group2 | CI | p-value | W | # Group1/<br># Group2 |
| Predicted transmission pairs | Case patient healthcare exposures | -0.00038 [-0.04788, 0.01271] | 0.8766 | 433 | 37/24 | -0.15521 [-0.27588, -0.05075] | 0.00010 | 12 | 12/16 | 0.04822 [0.00397, 0.07268] | 0.009657 | 390 | 27/20 |
| Predicted transmission pairs | Reference | 0.01335 [0.00916, 0.22699] | 2.542*10<sup>-9</sup> | 142144 | 37/4901 | 0.00224 [0.00052, 0.00604] | 0.01574 | 37168 | 12/4415 | 0.06766 [0.06532, 0.07038] | 1.339*10<sup>-10</sup> | 90864 | 27/3922 |
| Case patient healthcare exposures | Reference | 0.01455 [0.00637, 0.06647] | 1.417*10<sup>-5</sup> | 88976 | 24/4901 | 0.15671 [0.07015, 0.23174] | 1.478*10<sup>-9</sup> | 66205 | 16/4415 | 0.01631 [0.00639, 0.02173] | 2.887*10<sup>-6</sup> | 62972 | 20/3922 |

**Supplementary Table 3. Genome IDs and healthcare exposure data**

**Supplementary Figure 1.**
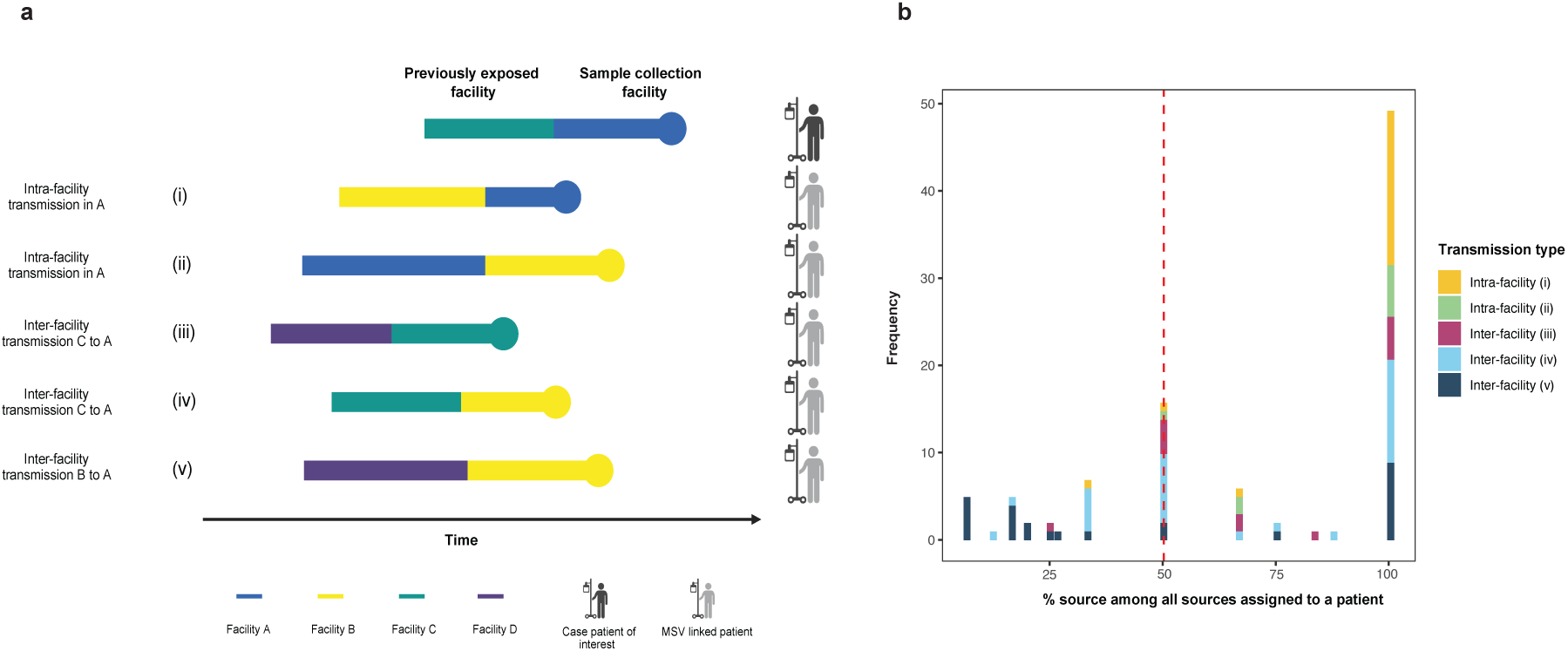
Transmission source prediction and most likely source identification. **(a)** Schematic of source facility assignment based on different types of shared healthcare exposures among MSV pairs. **(b)** The distribution of percent of shared exposures between case patients and their MSV linked pairs for each healthcare facility shared with a case patient and at least one MSV pair. If greater than 50% of shared exposures with MSV pairs were for a single source facility, then this was selected as a high-confidence source facility. If no facility was supported by greater than 50% of shared exposures with MSV pairs, then the source facility was deemed uncertain. 94% of cases had a single source facility supported by greater than 50% of the shared exposures to MSV-linked patients.

**Supplementary Figure 2.**
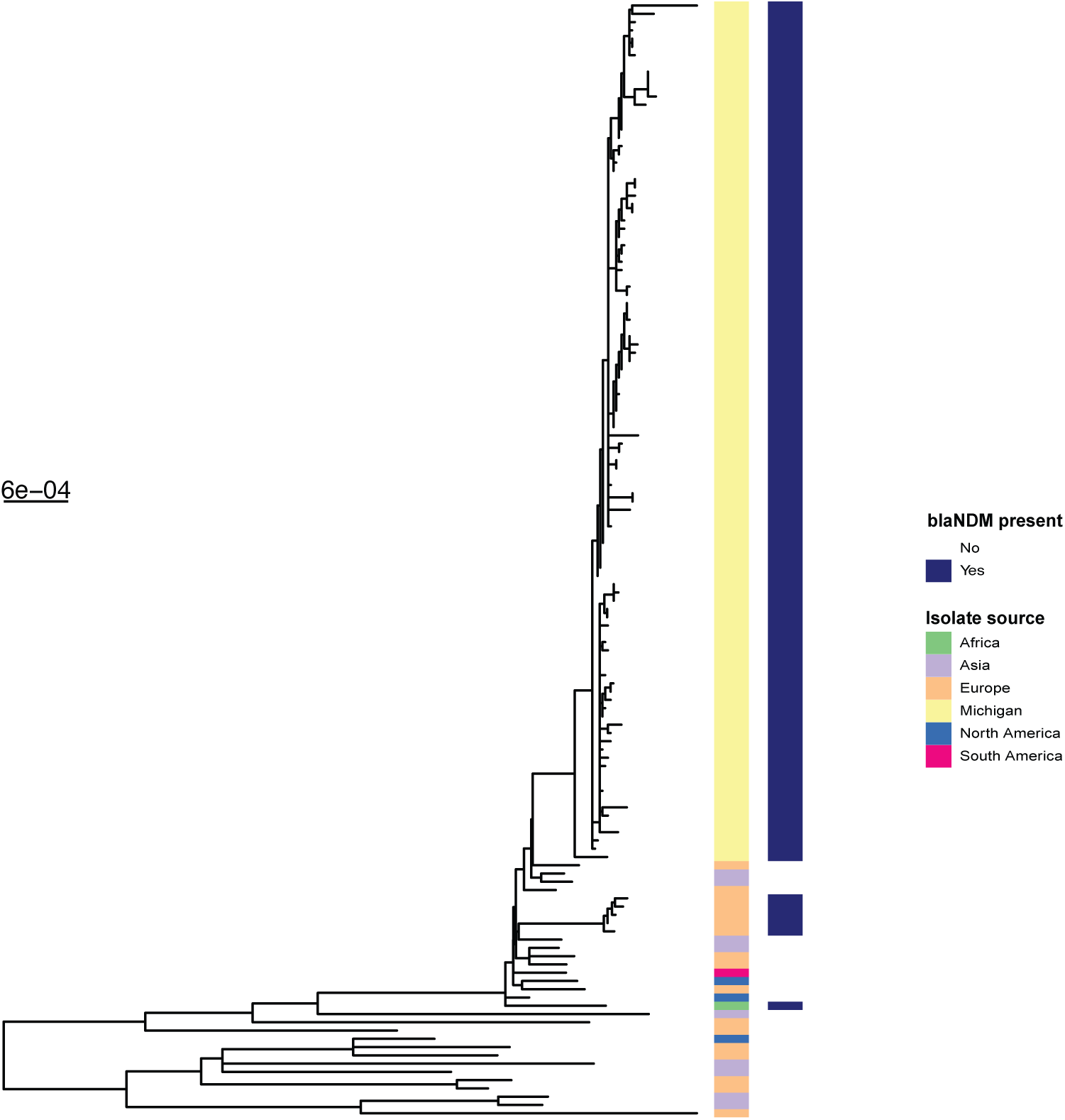
Whole-genome phylogeny including Michigan outbreak genomes and publicly available ST219 genomes. Midpoint rooted maximum likelihood tree of Michigan ST219 isolates and *K. pneumoniae* genomes downloaded from the PATRIC database that were within 200 SNVs of outbreak isolates.

**Supplementary Figure 3.**
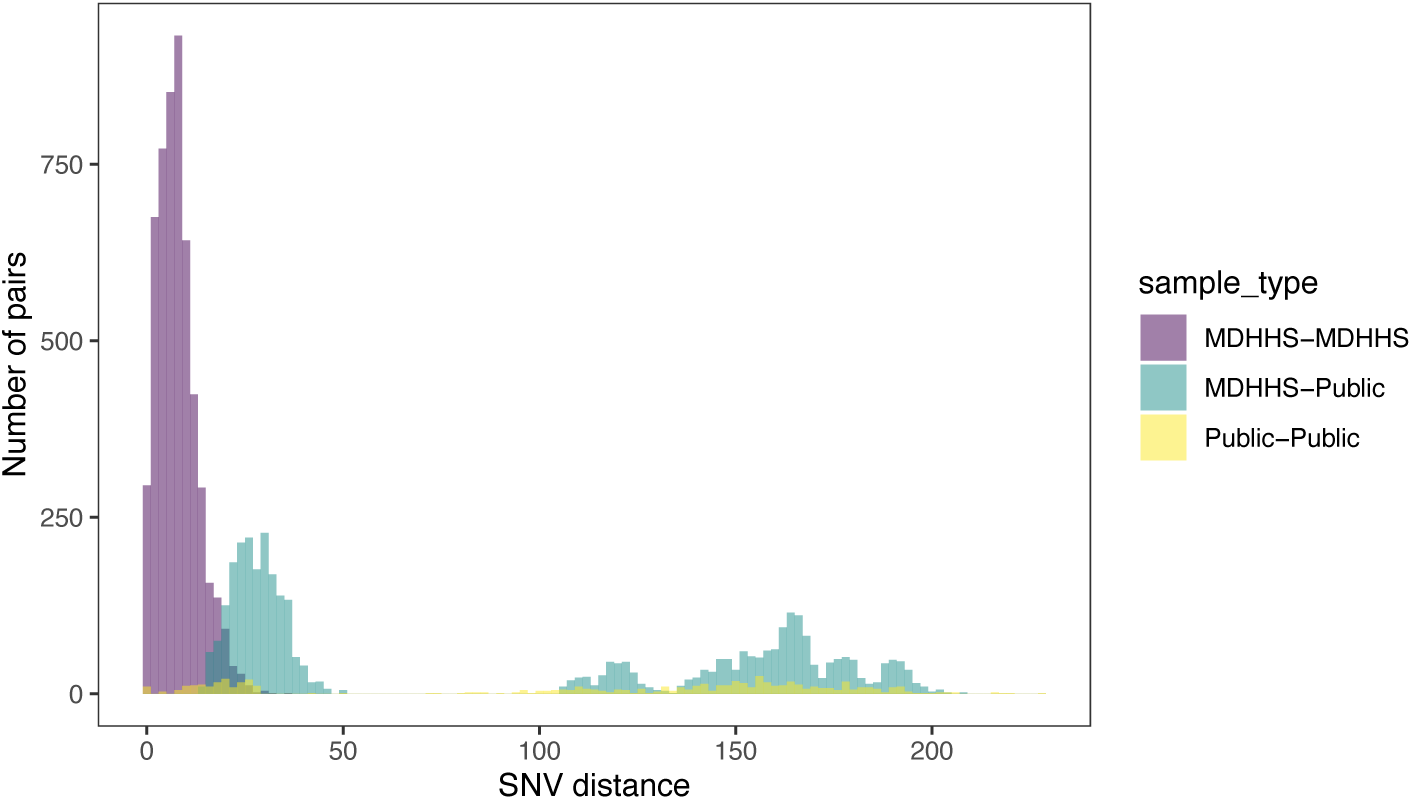
Pairwise distance histogram colored by pair types. Pairwise SNV distances are show for pairs of isolates collected in Michigan (purple), pairs of ST219 isolates downloaded from PATRIC (yellow) and pairs of isolates including one from Michigan and one from PATRIC (green).

**Supplementary Figure 4.**
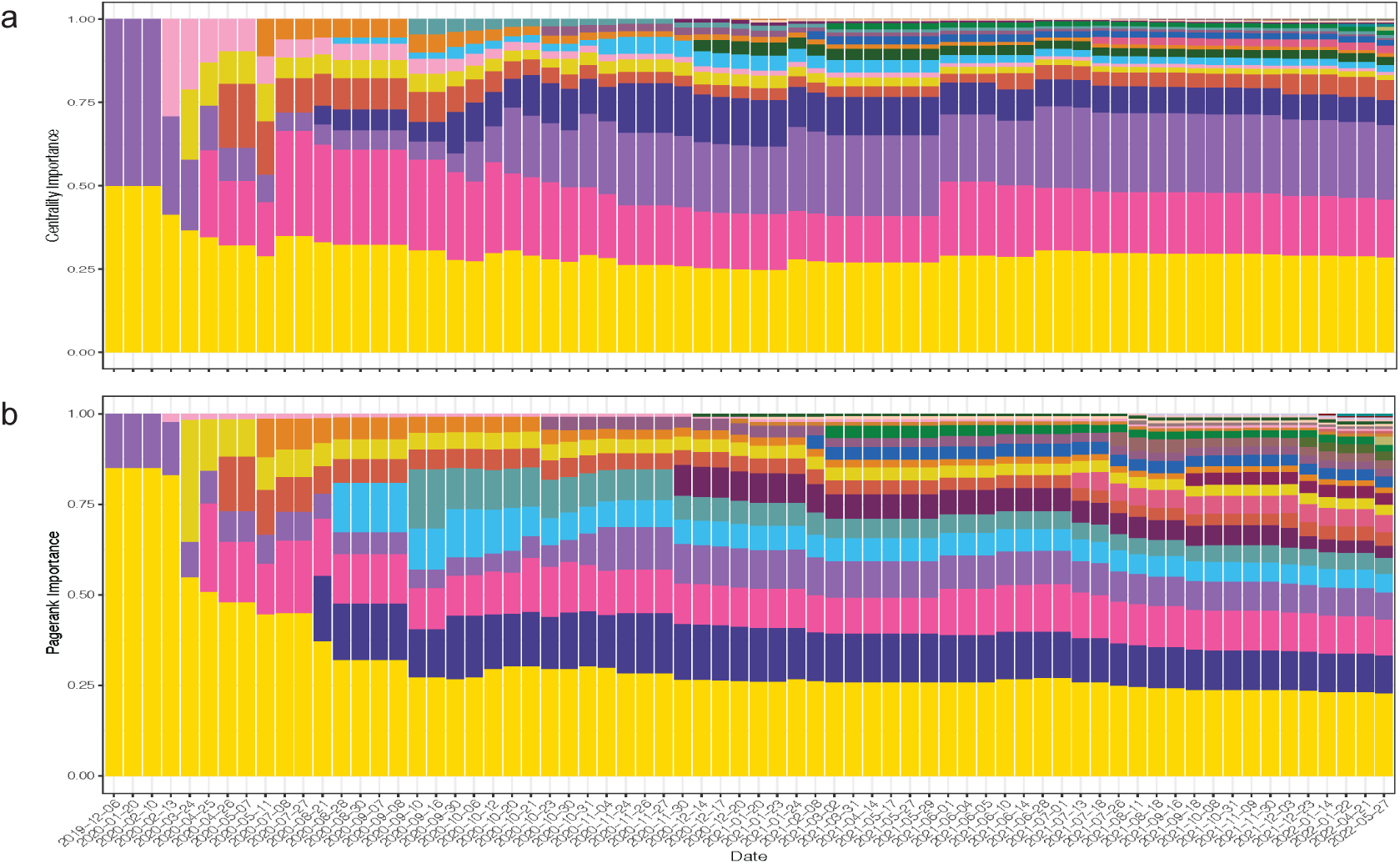
**(a)** Eigencentrality and **(b)** pagerank ranks indicating the relative importance of each facility in the predicted regional transmission network at timepoints when new isolates were collected.

**Supplementary Figure 5.**
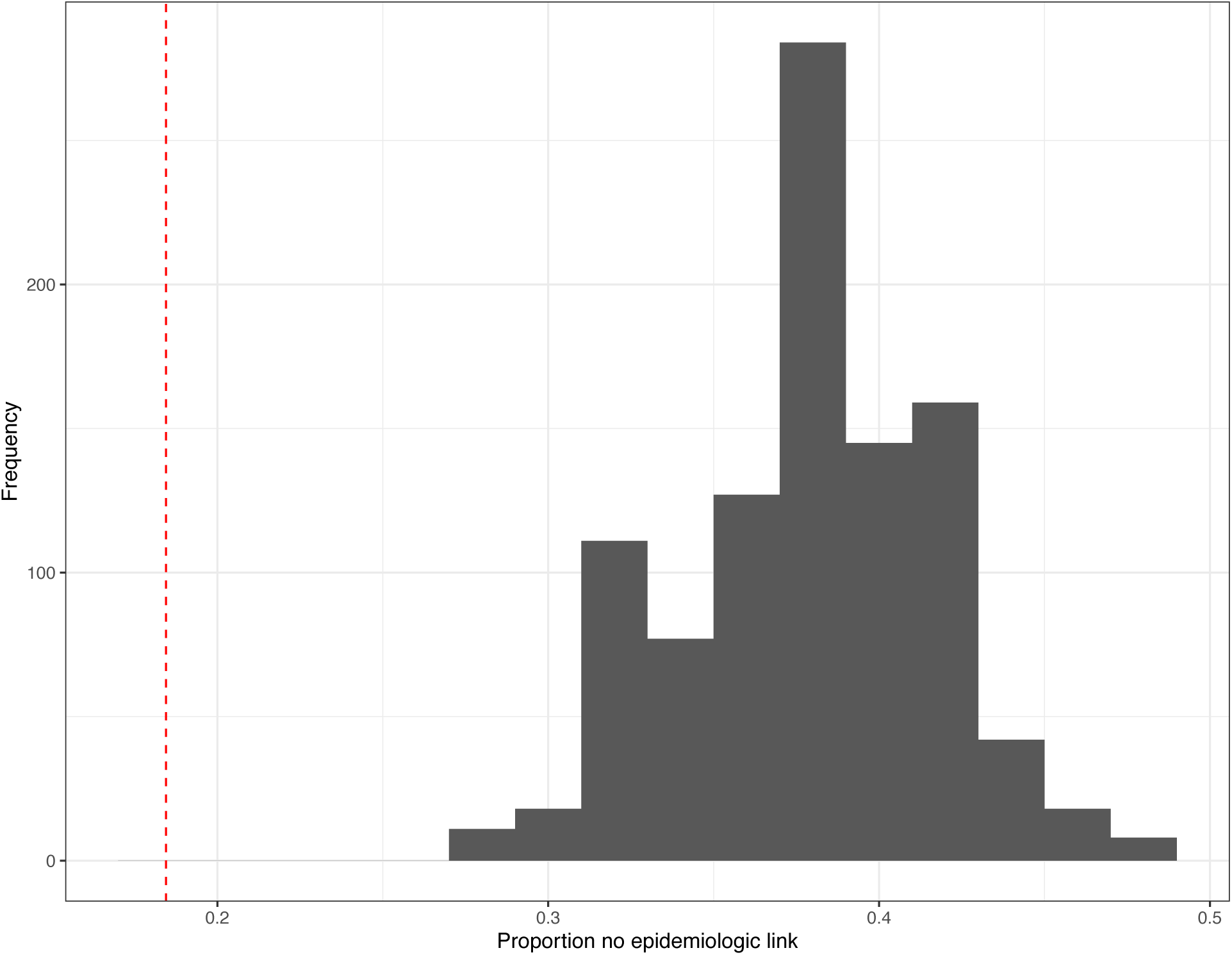
Proportion of MSV pairs without epidemiologic link in permutated dataset. The dashed link indicates the proportion of MSV pairs without epidemiologic links from the original dataset.

